# Associations of physical activity and sedentary time from childhood to adolescence with cognition in adolescence: The PANIC study

**DOI:** 10.1101/2025.06.04.25328954

**Authors:** Petri Jalanko, Marja H. Leppänen, Bert Bond, Jari A. Laukkanen, Timo A. Lakka, Eero A. Haapala

## Abstract

**Purpose:** Investigate the associations of cumulative exposure to physical activity (PA), sedentary time and screen time from childhood to adolescence over an 8-year follow-up with cognition in adolescence.

**Methods:** Altogether, 260 adolescents (136 boys) who were 15–17 years at 8-year follow-up were analysed. PA and sedentary time were assessed using an Actiheart®-device, and different types of PA, non-screen-based sedentary time and screen time were assessed by a questionnaire at baseline, 2-year and 8-year examinations. Cognition was assessed using CogState tests at 8-year examinations.

**Results:** Self-reported cumulative unsupervised PA from childhood to adolescence was inversely associated with accuracy in working memory task (standardized regression coefficient (β)=-0.127, 95% confidence interval (CI) −0.247 to −0.008) in adolescence. Self-reported non-screen-based sedentary time was inversely associated with reaction time (β=-0.176, 95% CI −0.299 to −0.053) and accuracy (β=-0.149, 95% CI −0.274 to −0.024) in working memory tasks. Self-reported screen time was inversely associated with reaction time (β=-0.194, 95% CI −0.316 to −0.071) and (β=-0.233, 95% CI −0.355 to −0.112) in both working memory tasks and directly associated with overall cognition (β=0.187, 95% CI 0.070-0.305).

**Conclusion:** Accumulating less unsupervised PA and more screen time from childhood to adolescence was associated with better cognition in adolescence.

## 1 Introduction

Insufficient levels of physical activity (PA) have been associated with poorer cognition in children ^1,2^ and adults ^3^. Childhood and adolescence are critical periods for cognitive and brain development ^4^, yet PA, particularly moderate to vigorous PA (MVPA), tends to decrease during this period, while sedentary time and screen time tend to increase ^5–7^. The effects of PA, along with prolonged sedentary time and screen time, on cognition may require prolonged cumulative exposure to low levels of PA and high levels of sedentary time and screen time, often beginning in childhood ^8^. Despite this, no evidence exists on the associations of cumulative exposure to PA, sedentary time and screen time from childhood to adolescence with cognition in adolescence ^9^. Moreover, little is known about the potential modifying role of sex in the association between PA and cognition in adolescents and the associations of PA from childhood to adolescence at different intensities with cognition in adolescents ^9,10^.

PA, sedentary time and screen time may influence cognition in adolescents by releasing neurotrophic factors and causing changes in brain structure and function, while also affecting sleep, stress, and anxiety ^11^. Nonetheless, the available cross-sectional evidence regarding the relationship between PA and cognition in adolescents remains mixed. Some cross-sectional studies suggest that device-assessed higher intensity PA ^10^, MVPA ^12^ and self-reported participation in organized sports ^13^ and active commuting to school ^14^ are directly associated with cognition in adolescents, whilst a previous study ^15^ suggests a weak, if any, association between self-reported extracurricular PA and cognition in children and adolescents. Furthermore, the respective evidence from longitudinal studies in adolescents is limited and inconsistent ^16,17^. Whilst an increase in device-assessed MVPA from the age of 9 to 15 years was associated with poorer working memory at 15 years ^16^, lower levels of self-reported extracurricular PA at the age of 6 years were associated with poorer cognition at 14 years ^17^. In summary, the existing evidence on the associations between PA and cognitive function in adolescents presents mixed findings, partly due to the different intensity and context of PA and the varying methods used to measure PA^15,16^. Therefore, further research that incorporates both self-reported and device-assessed PA is essential for a clearer understanding of how the intensity and context of PA influence the relationship between PA and cognitive function in adolescence.

Few studies have investigated the associations of sedentary time and screen time with cognition in adolescents ^16–18^. Higher self-reported sedentary time in mid-childhood was inversely associated with working memory in adolescence among boys ^17^. Moreover, changes in total self-reported screen time and video game playing from age 13 to 16 years were inversely associated with changes in working memory among girls ^18^. Wickel ^16^ found that children aged 9 years accumulating higher levels of device-assessed sedentary time had better overall cognition at 15 years. In addition, they found that a larger increase in device-assessed sedentary time between ages 9 to 15 years was beneficially associated with inhibition, working memory and overall cognition at the age of 15. This may be due to sedentary activities like reading, writing, doing homework and playing video and board games that can enhance cognition ^19,20^. However, Wickel ^16^ did not differentiate specific sedentary behaviours when studying the association between sedentary time and cognition in youth. In contrast, other researchers have found no association between self-reported sedentary time and cognition in adolescents ^13,14^. However, inaccuracies in reporting due to social desirability bias in self-reported measures may undermine the evidence in these studies ^21^. The existing evidence on the relationship between sedentary time and cognition is mixed, lacking device-assessed measurements of sedentary time and contextualization of sedentary activities.

Evidence for the modifying role of sex in the association of PA, sedentary time and screen time with cognition, as well as the associations of PA at different intensities with cognition in adolescents, remains unclear and limited ^10,12–16^. Differences in the associations between PA and cognition across various studies may be due to the distinct levels of physical, psychological, and brain development observed in boys and girls ^22,23^. The results of some studies indicate that sex does not modify the association of self-reported participation in organized sports ^13^ and extracurricular PA ^15^ with cognition in children and adolescents. However, some evidence suggests that the direct associations of device-assessed higher intensity PA ^10^ and self-reported active commuting to school ^14^ with cognition are stronger in adolescence among girls than boys. Moreover, studies on the role of PA intensity in these associations have found that only device-assessed MVPA ^12^ and higher PA intensity among adolescents ^10^ were favourably associated with cognition. These associations may be attributed to mechanisms that enhance cognitive function, including greater increases in BDNF, lactate, and neurovascular responses from MVPA and vigorous physical activity (VPA) compared to light physical activity (LPA) ^24–26^. In contrast, Wickel ^16^ found that increased levels of device-assessed LPA and MVPA from childhood to adolescence were inversely associated with working memory in adolescence. Identifying the potential moderating role of sex on the associations between PA and cognition, along with the associations of PA at varying intensities with cognition in adolescents, may assist in individualising PA guidelines.

The evidence on the longitudinal associations of PA, sedentary time and screen time with cognition from childhood to adolescence is limited and mixed. This is partly due to the varying contexts of PA and sedentary time, as well as the different methods used to assess them. Moreover, there have been no studies investigating the associations of cumulative exposure to PA, sedentary time and screen time from childhood to adolescence with cognition in adolescence ^16–18^ . Changes in cognition as a result of PA, sedentary time, and screen time may require long-term cumulative exposure to low levels of PA and high levels of sedentary time, often beginning in childhood ^8^. Therefore, we investigated the associations of cumulative PA, sedentary time and screen time from ages 8 to 16 years with cognition at age 16 using device-assessed and self-report measures. We also examined whether sex modifies these associations, and investigated how different intensities of PA from childhood to adolescence are associated with cognition in adolescence.

## 2 Methods

### 2.1 Study design and participants

The present analyses are based on data from the Physical Activity and Nutrition in Children (PANIC) study, which is an 8-year PA and dietary intervention study in a general population of children followed up until adolescence. For the present analysis, we treated the participants as a prospective cohort since the CogState cognitive test battery was administered only during the 8-year follow-up.

The Research Ethics Committee of the Hospital District of Northern Savo approved the study protocol in 2006 (Statement 69/2006) and 2015 (Statement 422/2015). Written informed consent was acquired from the parent or caregiver of each child, and every child provided assent to participation. The PANIC study was carried out following the principles of the Declaration of Helsinki, revised in 2008. The funding sources had no role in collecting, analyzing or interpreting the data or approving publications.

We invited 736 children 6–9 years of age who started the first grade in 16 primary schools of Kuopio (Figure 1). Altogether, 512 children (264 boys, 248 girls), who accounted for 70% of those invited, participated in the baseline examinations. The participants did not differ in age, sex or body mass index - standard deviation score (BMI-SDS) from all children who started the first grade in the city of Kuopio. We excluded six children from the study at baseline because of physical disabilities that could hamper intervention participation or lack of time or motivation to attend the study. As a result, data from 506 participants were used in the baseline statistical analyses. Altogether, 437 children (223 boys, 214 girls) who participated in the baseline examinations attended the 2-year follow-up examinations and represented 85% of those attending the baseline examinations.

**Figure 1.**
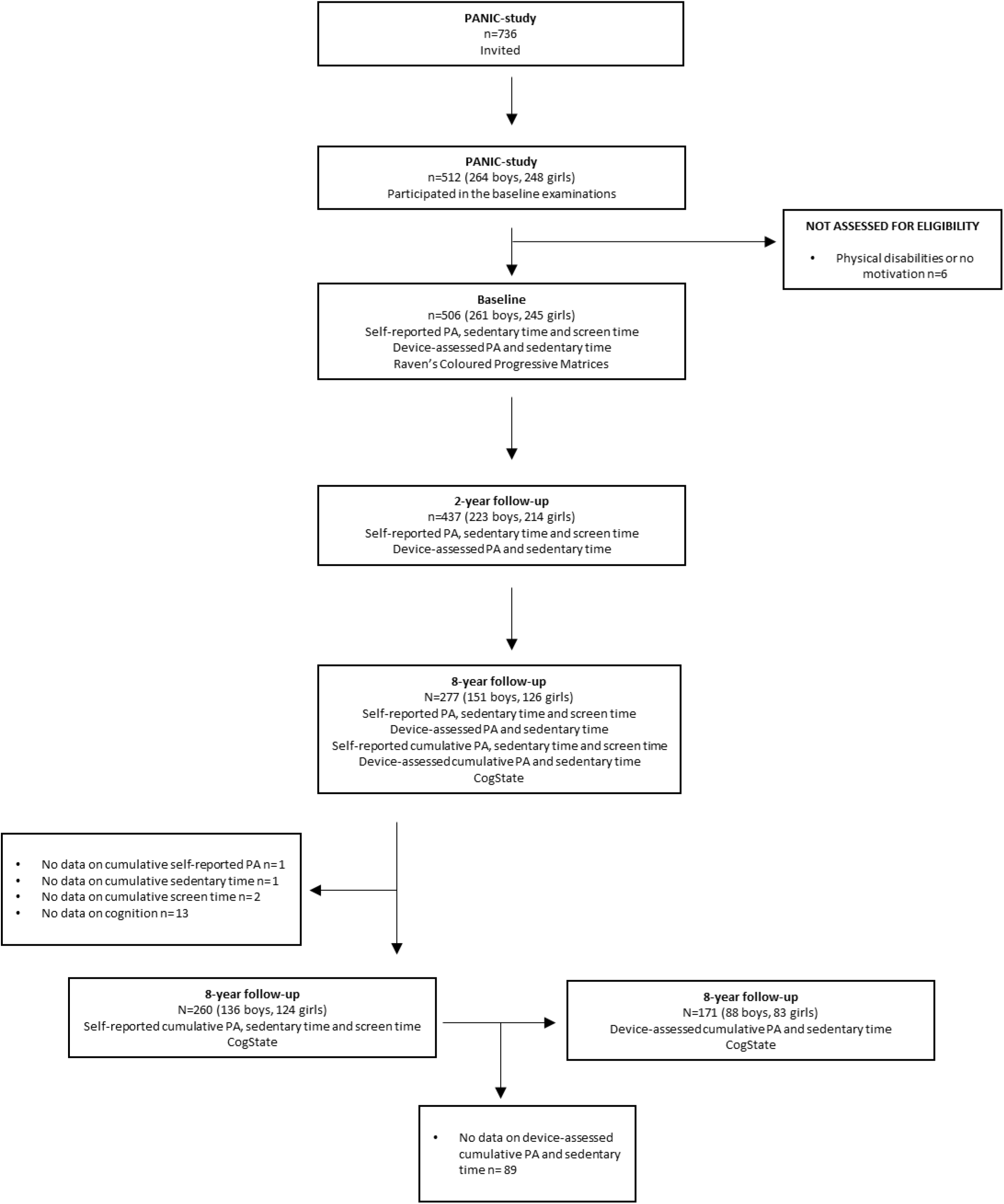
Study flow diagram. PA, Physical Activity

Finally, 277 adolescents (151 boys, 126 girls) participated in the 8-year follow-up examinations and represented 54% of those attending the baseline examinations. Those who participated in the 8-year follow-up examinations did not differ in age, BMI-SDS, the distribution of sex or study groups at baseline from those who dropped out. A total of 17 adolescents (15 boys; 2 girls) were excluded from the statistical analyses because they had no valid data on cumulative self-reported PA (n=1), sedentary time (n=1) screen time (n=2) or cognition (n=13) at the 8-year follow-up examinations. Therefore, 260 adolescents (136 boys; 124 girls) aged 15.8 ± 0.4 years from the 8-year follow-up examinations were used for the statistical analyses to explore the associations of self-reported cumulative PA, sedentary time and screen time with cognition. The included 136 boys had more advanced pubertal status (P=0.040) and lower levels of total PA (P=0.043) and self-reported unsupervised PA (P=0.013) than the 15 excluded boys. The 124 included girls had less self-reported screen time (P=0.044) than the two excluded girls. Altogether, 23 participants (13 boys; 10 girls) had no data on pubertal status. However, we included these participants in the statistical analyses because pubertal status was used only as a covariate. We replaced the missing data on pubertal status by multiple imputation using 10 imputed datasets. A total of 161 participants (88 boys; 73 girls) were included in the intervention group and 99 in the control group (48 boys; 51 girls). Of the 260 participants, 171 (88 boys; 83 girls) had data on device-assessed PA and sedentary time. These 171 participants were included in the statistical analyses to explore the associations of device-assessed cumulative PA and sedentary time with cognition. Of the 171 participants, a total of 105 participants (56 boys; 49 girls) were included in the intervention group and 66 in the control group (32 boys; 34 girls).

### 2.2 Assessment of body size and composition and pubertal status at 8-year follow-up

Two repeated measurements of body mass were carried out by the InBody^®^ 720 bioelectrical impedance device (Biospace, Seoul, South Korea) to an accuracy of 0.1 kg. The mean of these two values was used in the statistical analyses. Body stature was measured three times the adolescents standing in the Frankfurt plane without shoes using a wall-mounted stadiometer to an accuracy of 0.1 cm. The mean of the nearest two values was used in the statistical analyses. BMI was calculated by dividing body mass (kg) with stature squared (m^2^), and BMI-SDS was calculated using the Finnish references ^27^. Body fat percentage was measured using the Lunar^®^ dual-energy X-ray absorptiometry device (GE Medical Systems, Madison, WI, USA). A research physician assessed pubertal status using the five-stage scale described by Tanner ^28^. We used testicular volume assessed by an orchidometer to assess pubertal status in boys and breast development to assess pubertal status in girls ^28^.

### 2.3 Assessment of self-reported physical activity, non-screen-based sedentary time and screen time at baseline, 2-year follow-up, and 8-year follow-up

Self-reported PA, non-screen-based sedentary time and screen time were assessed using the PANIC Physical Activity Questionnaire administered by the parents at baseline and 2-year follow-up. At the 8-year follow-up, the adolescents completed the questionnaire themselves. The types of self-reported PA included unsupervised PA, organized sports (i.e., participation in organized sports training) and all organized exercise (e.g. sports, afterschool exercise clubs, and other supervised/coached exercise hobbies).

Non-screen-based sedentary time was calculated by summing up the time spent listening to music, playing a musical instrument, reading, writing, drawing, doing arts and crafts, playing board games, and resting. Screen time was calculated by summing up the time spent watching TV and videos, using a computer, playing video games, using a mobile phone, and playing mobile games. Sedentary behaviour and PA questionnaires with a similar structure to the PANIC Physical Activity Questionnaire, such as the Youth Physical Activity questionnaire, have shown good short–term repeatability over four days with an intraclass correlation of 0.86–0.92 ^29^.

### 2.4 Assessment of device-assessed physical activity and sedentary time at baseline, 2-year follow-up, and 8-year follow-up

PA and sedentary time were assessed using a combined heart rate and body movement sensor Actiheart^®^ (CamNtech Ltd., Papworth, UK) for a minimum of four consecutive days and analyzed in 60-second epochs at baseline, 2-year follow-up, and 8-year follow-up ^30^. The combined heart rate and movement sensor was attached to the child’s chest with two standard electrocardiographic electrodes (Bio Protech Inc., Donghwa-ri, South Korea). The children were asked to wear the monitor continuously, including sleep and water-based activities. Heart rate data were cleaned and individually calibrated using parameters obtained from the maximal cycle exercise test and were combined with movement sensor data to derive PA energy expenditure ^31^. Instantaneous PA energy expenditure was estimated using branched equation modelling and was expressed as time spent at intensity levels of standard metabolic equivalents (METs), one MET corresponding to 71.2 J/min/kg, in minutes per day ^30^. In the current study, vigorous physical activity (VPA) was defined as PAs at ≥7 METs, MVPA at ≥4 METs, LPA at > 1.5 and ≤ 4.0 METs and sedentary time at ≤ 1.5 METs, sleep excluded ^32^. These cut-off values are frequently used in studies examining PA among children and adolescents ^33^. TPA was calculated by summing LPA and MVPA (LPA + MVPA). PA data were defined valid if there was a minimum of 48 hours of activity recording in weekday and weekend day hours that included at least 12 hours from morning (3–9 am), midday (9 am–3 pm), afternoon (3–9 pm), and night (9 pm–3 am) to avoid potential bias from over-representing specific times and activities of the days.

### 2.5 Computation of the measures of device-assessed and self-reported cumulative PA, sedentary time and screen time

We used the area under the curve (AUC) approach for device-assessed and self-reported PA, sedentary time and screen time at baseline, 2-year follow-up, and 8-year follow-up to utilize all the data collected over the 8-year period and describe the the long-term exposure to PA, sedentary time and screen time from childhood to adolescence. The AUC method offers advantages over other longitudinal analysis models by measuring the long-term cumulative exposure, rather than solely analyzing changes in PA, sedentary time, and screen time, which can lead to biased causal-effect estimates ^34^. We estimated participant-specific curves for PA, sedentary time and screen time using an additive linear mixed model with cubic splines, which models non-linear effect as a random effect ^35,36^.

Y_ij = beta_0 + u_i + f(time_ij) + sex_i + e_ij, where f is the cubic spline, u_i the random intercept, beta_0 intercept, sex_i sex-effect, and e_ij error term. The modeling allowed the inclusion of a non-linear effect which was modeled by cubic spline in addition to random intercept for individuals. These AUC determined using the participant-specific curves were interpreted as long-term exposure to PA, sedentary time and screen time similar to Lai and colleagues.,^35^ and Hakala and colleagues.^37^

### 2.6 Assessment of cognition at baseline and 8-year follow-up

Psychomotor function, attention, working memory, paired-associate learning and overall cognition were assessed by a computerized CogState cognitive test battery (CogState Ltd, Melbourne, Australia) using a desktop personal computer ^38^ at the 8-year follow-up examinations. The construct validity of the CogState test battery has been demonstrated in a large group of healthy adults and children ^39^, and the test has been found to be applicable in different cultures since there are only minimal language requirements to undertake the tests ^40^.

The psychomotor function was assessed with the Detection Test. The participants were asked to click a button as quickly as possible when a playing card flipped over on a computer screen during the test. The test result was based on reaction time (in milliseconds), which was normalized using a log10 transformation. A lower score indicated better performance.

Attention was assessed by the Identification Task. The participants were asked to choose whether a revealed card was red or not during the task. The participants were required to press “yes” for the red card, and “no” for the black card. The test result was based on the reaction time (in milliseconds) and accuracy of each response. The reaction time was normalized using a log10 transformation. A lower score indicated better performance. The accuracy was normalized using an arcsine square-root transformation. A higher score indicated better performance.

Working memory was assessed by the One Back Task and Two Back Task. In the One Back Task, the participants were asked, using buttons “yes” or “no”, to answer whether the present card was the same as the previous card. In the Two Back Task, the participants were asked, using buttons “yes” or “no”, to answer whether a present card was the same as the card presented two cards ago. The test results were based on the reaction time (in milliseconds) and accuracy of each response. The reaction time was normalized using a log10 transformation. A lower score indicated better performance. The accuracy was normalized using an arcsine square-root transformation. A higher score indicated better performance.

Paired-associate learning was assessed by the Continuous Paired Associate Learning Task including two parts. First, the participants were asked to learn and remember abstract pictures and hidden patterns in different locations. Second, the participants were requested to recall where the re-displayed hidden picture had been located. The test score was the errors during all performance sequences, a lower score indicating better performance.

Overall cognition was calculated by taking the average of the standardized scores across all CogState tests. For the tests for which a lower score indicated better performance, the score was reversed (multiplied by −1) so that all outcome variables were in a uniform direction and a higher score indicated better performance.

Nonverbal reasoning was assessed using the Raven’s Coloured Progressive Matrices (RCPM) ^41^ at baseline and was used only as a covariate in the statistical analyses. The RCPM represents higher-order executive functioning involving inhibition, working memory and mental flexibility ^41^. The RCPM requires the ability to find similarities, differences, and discrete patterns and does not depend on acquired knowledge or language skills ^41^. The RCPM consist of three sets of 12 items. Each test page contains a large item or a pattern of items and six small items. The child was asked to select the correct small item, which completes the large item or the set of items. The RCPM score was the number of correct answers, ranging from 0 to 36, with higher scores indicating better cognitive performance.

### 2.7 Statistical analyses

All statistical analyses were carried out using the SPSS statistical analysis software, Version 27.0 (IBM Corp., Armonk, NY, USA). The normality of the distributions of the variables was tested with the Kolmogorov–Smirnov test and visually from histograms. Differences in descriptive characteristics, PA, sedentary time and screen time between sexes were analyzed by the Student’s T-test, the Mann Whitney U-test or the Chi-square Test. The associations of cumulative PA, sedentary time and screen time with cognition were investigated using linear regression analyses adjusted for age, sex and parental education at 8-year follow-up and the

RCPM score at baseline. The data were adjusted for parental education as it has been associated directly with cognition ^42^ and PA ^43^. Age, sex, parental education and the RCPM score were entered into the linear regression models in the first step, and the measures of cumulative PA, sedentary time and screen time were entered separately into the models in the second step. The standardised regression coefficients β and their 95% confidence intervals with the corresponding *P*-values were reported for each factor. Differences and associations with *P*-values ≤ 0.05 were considered statistically significant. The data were corrected for multiple comparisons using the Benjamini-Hochberg false discovery rate (FDR) with an FDR value of 0.2 (FDR_0.2_). If the associations of cumulative PA, sedentary time and screen time with cognition were statistically significant after adjustment for sex, age and parental education at 8-year follow-up and the RCPM score at baseline, the data were additionally adjusted for body fat percentage and pubertal status at 8-year follow-up and study group. Body fat percentage, pubertal status and study group were entered into the models separately to evaluate their independent influence on the associations. Analyses by sex were conducted similarly, except for not using sex as a covariate. Finally, we investigated whether sex modified the associations of cumulative PA, sedentary time and screen time with cognition using general linear models.

## 3 Results

### 3.1 Characteristics of participants

Boys were heavier and taller and had lower body fat percentage and their parents had a higher level of education than girls’ parents at the 8-year follow-up. However, girls had more advanced pubertal status than boys (Table 1). Boys accumulated more self-reported total PA, unsupervised PA, organised sports and screen time and less self-reported non-screen-based sedentary time than girls. In addition, boys accumulated more device-assessed MVPA and VPA than girls. Boys had shorter reaction times in the Detection Test, One Back Task and Two

**Table 1.** Descriptive characteristics of all 260 participants at 8-year follow-up.

|  | All adolescents (n<br>=260) | Boys (n =136) | Girls (n =124) | P-value |
| --- | --- | --- | --- | --- |
| **Background characteristics** |  |  |  |  |
| Age (years) | 15.8 ± 0.4 | 15.8 ± 0.4 | 15.8 ± 0.4 | 0.349 |
| Body mass (kg) | 62.0 ± 12.5 | 65.7 ± 13.9 | 57.9 ± 9.2 | **<0.001** |
| Body stature (cm) | 171.3 ± 8.6 | 176.4 ± 7.4 | 165.7 ± 5.8 | **<0.001** |
| Body mass index - standard deviation score | 0.0 ± 1.0 | -0.1 ± 1.1 | 0.1 ± 0.9 | 0.262 |
| Body fat percentage (%) | 22.7 ± 10.2 | 17.1 ± 9.4 | 28.9 ± 7.1 | **<0.001** |
| Pubertal status, n (%) † |  |  |  | **0.004** |
| Stage 3 | 19 (8.0) | 15 (12.2) | 4 (3.5) |  |
| Stage 4 | 137 (57.8) | 76 (61.8) | 61 (53.5) |  |
| Stage 5 | 81 (34.2) | 32 (26.0) | 49 (43.0) |  |
| Parental education (%) |  |  |  | **0.026** |
| Vocational school or less | 13.4 | 13.2 | 13.7 |  |
| Polytechnic | 40.4 | 33.1 | 48.4 |  |
| University degree | 46.2 | 53.7 | 37.9 |  |
| **Self-reported PA and sedentary behaviour** |  |  |  |  |
| Total physical activity (min/d) | 147.9 ± 106.3 | 172.6 ± 126.5 | 120.9 ± 69.5 | **<0.001** |
| Organised sports (min/d) | 26.8 ± 35.0 | 32.6 ± 39.9 | 20.5 ± 28.9 | **0.033** |
| All organised exercise (min/d) | 33.9 ± 42.6 | 40.8 ± 49.5 | 26.3 ± 31.9 | 0.221 |
| Unsupervised physical activity (min/d) | 86.1 ± 85.2 | 101.1 ± 97.4 | 69.7 ± 66.0 | **0.004** |
| Non-screen-based sedentary time (min/d) | 192.1 ± 162.5 | 154.9 ± 130.4 | 232 ± 183.8 | **<0.001** |
| Screen time (min/d) | 345.1 ± 191.6 | 375.9 ± 208.6 | 311.3 ± 165.4 | **0.002** |
| **Device-assessed PA and sedentary time** |  |  |  |  |
| Total physical activity (min/d) § | 367.0 ± 133 | 376.6 ± 133.8 | 352.4 ± 132.1 | 0.368 |
| Light physical activity (min/d) § | 319.9 ± 113.3 | 323.1 ± 111.9 | 315.1 ± 116.6 | 0.727 |
| Moderate-to-vigorous physical activity (min/d) § | 47.1 ± 36.6 | 53.6 ± 39.8 | 37.4 ± 28.8 | **0.044** |
| Vigorous physical activity (min/d) § | 11.4 ± 14.4 | 14.7 ± 16.1 | 6.5 ± 9.6 | **0.013** |
| Sedentary time (min/d) § | 607 ± 134.7 | 603.3 ± 137.1 | 613.5 ± 132.4 | 0.709 |
| **Cognition** |  |  |  |  |
| Overall cognition (standardized score) | 0.00 ± 0.51 | 0.02 ± 0.49 | -0.01 ± 0.52 | 0.447 |
| Detection Test (log10msec) # | 2.50 ± 0.06 | 2.49 ± 0.05 | 2.51 ± 0.06 | **0.031** |
| Identification Task (log10msec) # | 2.68 ± 0.06 | 2.68 ± 0.06 | 2.68 ± 0.05 | 0.840 |
| One Back Task (log10msec) # | 2.87 ± 0.10 | 2.85 ± 0.09 | 2.89 ± 0.10 | **0.004** |
| Two Back Task (log10msec) # | 2.96 ± 0.11 | 2.93 ± 0.10 | 2.99 ± 0.10 | **< 0.001** |
| Identification Task (arcsine proportion correct) ㉔ | 1.41 ± 0.15 | 1.38 ± 0.16 | 1.43 ± 0.13 | **0.007** |
| One Back Task (arcsine proportion correct) $\pi$ | 1.39 ± 0.15 | 1.38 ± 0.14 | 1.39 ± 0.17 | 0.217 |
| Two Back Task (arcsine proportion correct) $\pi$ | 1.27 ± 0.18 | 1.28 ± 0.17 | 1.25 ± 0.18 | 0.208 |
| Continuous Paired Associate Learning Task (errors) | 31 ± 33 | 38 ± 38 | 23 ± 24 | **< 0.001** |

The data are means (SDs) and corresponding *P*-values from the Student's T-test for independent samples for continuous variables with normal distributions, from the Mann–Whitney U-test for continuous variables with skewed distributions and the Chi-square Test for categorical variables. *P*-values indicating statistically significant differences are bolded.
† 237 adolescents (123 boys, 114 girls); § 103 adolescents (62 boys; 41 girls): # mean of the log10 transformed reaction times for correct responses, lower score = better performance $\pi$ arcsine transformation of the square root of the proportion of correct responses, higher score = better performance. Abbreviations: PA, physical activity.

Back Task, poorer accuracy in the Identification Test and more errors in the Continuous Paired Associate Learning Task than girls.

### 3.2 Associations of cumulative physical activity and sedentary time with cognition

Higher levels of self-reported cumulative unsupervised PA were associated with poorer accuracy in the Two Back Task after adjustment for sex, age and parental education at 8-year follow-up and the RCPM score at baseline (Table 2). However, the association did not remain statistically significant after FDR_0.2_ correction. Higher levels of self-reported cumulative non-screen-based sedentary time were associated with shorter reaction time in the Two Back Task and poorer accuracy in the One Back Task (Table 2). Higher levels of self-reported cumulative screen time were associated with shorter reaction time in the One Back Task and Two Back Task and better overall cognition (Table 2). These associations remained statistically significant after FDR_0.2_ correction. After further adjustment for body fat percentage and pubertal status at 8-year follow-up or study group, all these associations remained materially unchanged. Other self-reported measures of PA were not associated with cognition. Device-assessed cumulative PA and sedentary time were not associated with cognition (Table 3).

**Table 2.** Associations of self-reported cumulative PA, sedentary time and screen time with measures of cognition in all 260 participants.

|  | Detection Test<br>(reaction time) |  | Identification Test<br>(reaction time) |  | One Back Task<br>(reaction time) |  | Two Back Task<br>(reaction time) |  | Identification Test<br>(accuracy) |  | One Back Task<br>(accuracy) |  | Two Back Task<br>(accuracy) |  | Continuous Paired<br>Associate Learning<br>Test<br>(errors) |  | Overall cognition |  |
| --- | --- | --- | --- | --- | --- | --- | --- | --- | --- | --- | --- | --- | --- | --- | --- | --- | --- | --- |
| | $\beta$<br>(95% CI) | p | $\beta$<br>(95% CI) | p | $\beta$<br>(95% CI) | p | $\beta$<br>(95% CI) | p | $\beta$<br>(95% CI) | p | $\beta$<br>(95% CI) | p | $\beta$<br>(95% CI) | p | $\beta$<br>(95% CI) | p | $\beta$<br>(95% CI) | p |
| **Total PA** | -0.075<br>(-0.198,<br>0.048) | 0.232 | -0.069<br>(-0.197,<br>0.059) | 0.290 | -0.036<br>(-0.162,<br>0.090) | 0.572 | 0.019<br>(-0.107,<br>0.145) | 0.765 | -0.001<br>(-0.129,<br>0.127) | 0.987 | 0.089<br>(-0.037,<br>0.215) | 0.165 | -0.079<br>(-0.200,<br>0.042) | 0.199 | 0.045<br>(-0.077,<br>0.166) | 0.470 | -0.007<br>(-0.128,<br>0.114) | 0.908 |
| **All**<br>**organised**<br>**exercise** | -0.076<br>(-0.195,<br>0.043) | 0.207 | -0.120<br>(-0.243,<br>0.003) | 0.056 | -0.071<br>(-0.192,<br>0.050) | 0.249 | -0.027<br>(-0.148,<br>0.095) | 0.668 | 0.005<br>(-0.119,<br>0.129) | 0.935 | 0.043<br>(-0.080,<br>0.165) | 0.491 | 0.002<br>(-0.115,<br>0.119) | 0.970 | 0.031<br>(-0.087,<br>0.148) | 0.606 | 0.048<br>(-0.069,<br>0.164) | 0.422 |
| **Unsupervis**<br>**ed PA** | -0.038<br>(-0.160,<br>0.085) | 0.547 | -0.002<br>(-0.129,<br>0.126) | 0.980 | 0.002<br>(-0.123,<br>0.127) | 0.976 | 0.040<br>(-0.085,<br>0.165) | 0.531 | -0.005<br>(-0.132,<br>0.123) | 0.944 | 0.078<br>(-0.048,<br>0.204) | 0.223 | **-0.127**<br>**(-0.247,**<br>**-0.008)** | **0.037\*** | 0.043<br>(-0.078,<br>0.164) | 0.484 | -0.054<br>(-0.174,<br>0.067) | 0.381 |
| **Organised**<br>**sports** | -0.071<br>(-0.190,<br>0.048) | 0.238 | -0.121<br>(-0.244,<br>0.002) | 0.053 | -0.081<br>(-0.202,<br>0.040) | 0.189 | -0.031<br>(-0.153,<br>0.091) | 0.615 | -0.018<br>(-0.142,<br>0.106) | 0.779 | 0.037<br>(-0.086,<br>0.159) | 0.557 | 0.027<br>(-0.090,<br>0.144) | 0.651 | 0.035<br>(-0.083,<br>0.152) | 0.561 | 0.047<br>(-0.070,<br>0.164) | 0.428 |
| **Non-**<br>**screen-**<br>**based**<br>**sedentary**<br>**time** | -0.030<br>(-0.153,<br>0.092) | 0.628 | -0.121<br>(-0.248,<br>0.006) | 0.061 | -0.114<br>(-0.239,<br>0.010) | 0.071 | **-0.176**<br>**(-0.299,**<br>**-0.053)** | **0.005\*** | 0.022<br>(-0.106,<br>0.149) | 0.737 | **-0.149**<br>**(-0.274,**<br>**-0.024)** | **0.019\*** | -0.059<br>(-0.179,<br>-0.061) | 0.333 | 0.035<br>(-0.086,<br>0.156) | 0.569 | 0.053<br>(-0.067,<br>0.173) | 0.388 |
| **Screen time** | -0.043<br>(-0.165,<br>0.079) | 0.490 | -0.094<br>(-0.221,<br>0.032) | 0.143 | **-0.194**<br>**(-0.316,**<br>**-0.071)** | **0.002\*** | **-0.233**<br>**(-0.355,**<br>**-0.112)** | **<0.001**<br>**\*\*\*** | 0.036<br>(-0.091,<br>0.163) | 0.576 | 0.037<br>(-0.088,<br>0.163) | 0.560 | 0.029<br>(-0.091,<br>0.149) | 0.632 | -0.028<br>(-0.148,<br>0.092) | 0.646 | **0.187**<br>**(0.070,**<br>**0.305)** | **0.002\***<br>**\*** |

The data are standardised regression coefficients $\beta$ with their 95% confidence intervals (95% CIs) and *P*-values from linear regression analyses adjusted for sex, age, and parental education at 8-year follow-up and the Raven's Coloured Progressive Matrices score at baseline. \**P*<0.05; \*\**P*<0.01; \*\*\* *P*<0.001.
Abbreviations: PA, physical activity.

**Table 3.** Associations of device-assessed cumulative PA and sedentary time with measures of cognition in 171 participants.

|  | Detection Test<br>(reaction time) |  | Identification Test<br>(reaction time) |  | One Back Task<br>(reaction time) |  | Two Back Task<br>(reaction time) |  | Identification Test<br>(accuracy) |  | One Back Task<br>(accuracy) |  | Two Back Task<br>(accuracy) |  | Continuous Paired<br>Associate Learning<br>Test<br>(errors) |  | Overall cognition |  |
| --- | --- | --- | --- | --- | --- | --- | --- | --- | --- | --- | --- | --- | --- | --- | --- | --- | --- | --- |
| | $\beta$<br>(95% CI) | p | $\beta$<br>(95% CI) | p | $\beta$<br>(95% CI) | p | $\beta$<br>(95% CI) | p | $\beta$<br>(95% CI) | p | $\beta$<br>(95% CI) | p | $\beta$<br>(95% CI) | p | $\beta$<br>(95% CI) | p | $\beta$<br>(95% CI) | p |
| **TPA** | -0.092<br>(-0.267,<br>0.083) | 0.300 | -0.086<br>(-0.271,<br>0.098) | 0.358 | -0.035<br>(-0.215,<br>0.146) | 0.705 | 0.057<br>(-0.125,<br>0.240) | 0.534 | 0.005<br>(-0.180,<br>0.190) | 0.957 | 0.035<br>(-0.146,<br>0.216) | 0.707 | -0.140<br>(-0.312,<br>0.033) | 0.112 | -0.026<br>(-0.201,<br>0.150) | 0.774 | -0.017<br>(-0.193,<br>0.158) | 0.845 |
| **LPA** | -0.027<br>(-0.186,<br>0.131) | 0.734 | 0.059<br>(-0.108,<br>0.227) | 0.486 | 0.046<br>(-0.117,<br>0.209) | 0.579 | 0.038<br>(-0.127,<br>0.203) | 0.650 | -0.047<br>(-0.214,<br>0.121) | 0.583 | 0.077<br>(-0.086,<br>0.241) | 0.352 | 0.000<br>(-0.158,<br>0.157) | 0.997 | 0.098<br>(-0.060,<br>0.257) | 0.223 | -0.061<br>(-0.220,<br>0.098) | 0.450 |
| **MVPA** | -0.067<br>(-0.268,<br>0.133) | 0.509 | -0.066<br>(-0.278,<br>0.145) | 0.536 | -0.176<br>(-0.381,<br>0.029) | 0.092 | -0.091<br>(-0.299,<br>0.118) | 0.392 | -0.017<br>(-0.228,<br>0.195) | 0.877 | 0.084<br>(-0.123,<br>0.291) | 0.423 | 0.144<br>(-0.054,<br>0.342) | 0.152 | -0.075<br>(-0.276,<br>0.126) | 0.462 | 0.157<br>(-0.043,<br>0.356) | 0.123 |
| **VPA** | -0.049<br>(-0.251,<br>0.153) | 0.634 | -0.132<br>(-0.344,<br>0.081) | 0.222 | -0.073<br>(-0.281,<br>0.135) | 0.488 | -0.020<br>(-0.230,<br>0.190) | 0.851 | 0.023<br>(-0.190,<br>0.236) | 0.833 | 0.079<br>(-0.129,<br>0.287) | 0.455 | 0.109<br>(-0.090,<br>0.309) | 0.281 | 0.052<br>(-0.150,<br>0.255) | 0.610 | 0.074<br>(-0.128,<br>0.276) | 0.471 |
| **Sedentary**<br>**time** | 0.043<br>(-0.104,<br>0.191) | 0.562 | -0.028<br>(-0.184,<br>0.127) | 0.721 | -0.027<br>(-0.179,<br>0.124) | 0.722 | -0.049<br>(-0.202,<br>0.104) | 0.528 | 0.016<br>(-0.139,<br>0.171) | 0.841 | -0.032<br>(-0.184,<br>0.120) | 0.682 | -0.064<br>(-0.209,<br>0.082) | 0.390 | -0.047<br>(-0.195,<br>0.100) | 0.527 | 0.021<br>(-0.127,<br>0.168) | 0.782 |

The data are standardised regression coefficients $\beta$ and their 95% confidence intervals (95% CIs) and *P*-values from linear regression analyses adjusted for sex, age and parental education at 8-year follow-up and the Raven's Coloured Progressive Matrices score at baseline.
Abbreviations: TPA total physical activity; LPA, light physical activity; MVPA, moderate-to-vigorous physical activity; VPA, vigorous physical activity

### 3.3 Associations of cumulative physical activity and sedentary time with cognition in boys

Self-reported cumulative unsupervised PA was inversely associated with accuracy in the Two Back Task in boys after adjustment for age and parental education at 8-year follow-up and the RCPM score at baseline (Supplementary Table 1). Higher levels of self-reported cumulative organised sports were associated with shorter reaction time in the One Back Task. Higher levels of self-reported cumulative non-screen-based sedentary time were associated with shorter reaction time in the Two Back Task. Higher levels of self-reported cumulative screen time were associated with shorter reaction time in the Two Back Task. Device-assessed cumulative PA and sedentary time were not associated with cognition in boys (Supplementary Table 2). All these associations remained after FDR_0.2_ correction, except the inverse association between cumulative organised sports participation and reaction time in the One Back Task. All these associations also remained materially unchanged after further adjustment for body fat percentage, pubertal status or study group.

### 3.4 Associations of cumulative physical activity and sedentary time with cognition in girls

Higher levels of self-reported cumulative screen time were associated with shorter reaction times in the One Back Task in girls (Supplementary Table 3). Higher levels of self-reported cumulative screen time were associated with shorter reaction times in the Two Back Task and better overall cognition. Higher levels of device-assessed cumulative LPA were associated with better accuracy in the One Back Task (Supplementary Table 4). All these associations remained after FDR_0.2_ correction. All these associations also remained materially unchanged after further adjustment for body fat percentage, pubertal status or study group.

### 3.5 Sex differences in the associations of cumulative physical activity and sedentary time with cognition

Higher levels of device-assessed cumulative LPA were associated with better accuracy in the One Back Task test in girls (β 0.269, 95% CI 0.057 to 0.480) but not in boys (β −0.119, 95% CI −0.331 to 0.093; *P* 0.025 for interaction. Moreover, the associations between device-assessed cumulative LPA and reaction time in the Two Back Task in boys (β −0.141, 95% CI−0.363 to 0.081) and girls (β 0.182, 95% CI −0.042 to 0.405; *P*=0.048 for interaction) and between MVPA and accuracy in the Identification Test in boys (β −0.096, 95% CI −0.331 to 0.138) and girls (β 0.215, 95% CI −0.009 to 0.439; *P*=0.047 for interaction) were different (Supplementary Table 5). However, these associations between PA and cognition were statistically non-significant. Finally, higher levels of self-reported cumulative screen time were associated with shorter reaction times in the One Back Task in girls (β −0.335, 95% CI −0.503 to −0.167) but not in boys (β −0.069, 95% CI −0.224 to 0.196; *P* 0.008 for interaction) (Supplementary Table 6). No other sex differences in the associations of cumulative PA or sedentary time with cognition were observed (Supplementary Tables 5 and 6).

## 4. Discussion

To the best of the authors’ knowledge, this study is the first to investigate the associations of cumulative exposure to PA, sedentary time, and screen time from childhood to adolescence with cognition in adolescence using both device-assessed and self-reported measures. We found that higher levels of self-reported cumulative unsupervised PA from childhood to adolescence were associated with poorer working memory in adolescence. Higher levels of self-reported cumulative non-screen-based sedentary time from childhood to adolescence were associated with shorter reaction times but poorer accuracy in the working memory task in adolescence. Moreover, higher levels of self-reported cumulative screen time from childhood to adolescence were associated with better working memory and overall cognition in adolescence. Device-assessed cumulative PA or sedentary time was not associated with cognition in the whole study population. Higher levels of self-reported cumulative organised sports and non-screen-based sedentary time from childhood to adolescence were associated with better working memory among boys but not among girls. In addition, higher levels of self-reported cumulative unsupervised PA from childhood to adolescence were related to poorer working memory in adolescence among boys but not among girls. Higher levels of device-assessed cumulative LPA from childhood to adolescence were associated with better working memory in adolescence among girls but not among boys. Finally, higher levels of self-reported cumulative screen time from childhood to adolescence were associated with better overall cognition in adolescence among girls but not among boys.

The inverse association between self-reported cumulative unsupervised PA from childhood to adolescence and working memory in adolescence contradicts the results of previous studies showing direct cross-sectional ^13,14^ and longitudinal ^17^ associations of self-reported organized sports ^13^, active commuting to school ^14^ and extracurricular PA ^17^ with cognition in adolescents. However, whereas Wickel ^16^ reported an inverse association between a change in device-assessed MVPA from the age of 9 years to the age of 15 years and working memory at the age of 15 years, we found that cumulative device-assessed MVPA was not associated with cognition in adolescents. Therefore, these inconsistent findings suggest that higher levels of PA in some circumstances may be related to poorer cognition, but that the associations may depend on several other factors, such as population characteristics, the context of PA or the method used to assess PA. One explanation for our observations may be that unsupervised PA replaces certain activities that may enhance cognition, such as reading and doing homework. Inconsistencies between self-reported and device-assessed PA and sedentary time and their associations with cognition may be due to inaccurate reporting of PA and sedentary time. The reasons for the reporting inaccuracy may be social desirability bias or parents’ unawareness of their children’s PA and sedentary time when they are not with them ^21^. Additionally, the lack of significant associations between device-assessed PA and cognition might result from limited variability in MVPA in adolescents, which can mask small effects. Furthermore, device-assessed measurements of PA may overlook important qualitative and contextual aspects of PA ^13,15,17^. While the exact reasons for the inverse association between self-reported cumulative unsupervised PA and working memory are unknown, it is important to note that this association needs to be interpreted cautiously, as the inverse association did not remain after FDR correction and other measures of cumulative PA were not associated with cognition.

We found direct associations of self-reported cumulative non-screen-based sedentary time and screen time from childhood to adolescence with cognition in adolescence. Nevertheless, we did not observe an association between device-assessed cumulative sedentary time from childhood to adolescence and cognition in adolescence, contradicting the results of Wickel ^16^, who reported that increased device-assessed sedentary time from the age of 9 to 15 years was associated with better cognition at the age of 15 years. However, the evidence on the association between device-assessed sedentary time and cognition in youth is mixed and from cross-sectional studies ^44,45^. One explanation for our finding may be that non-screen-based sedentary time and screen time include activities that may improve cognition, such as reading, writing, doing homework and playing video and board games ^19,20^. The lack of significant associations between device-assessed sedentary time and cognition in our study aligns with previous systematic review and may result from context-dependence not captured by device-assessed measurements ^45^. Additionally, using a combined accelerometer and heart rate monitor placed on the chest, rather than on the thigh, may lead to an underestimation of the time spent in sedentary activities, as the thigh placement would provide a more accurate estimate of sedentary time ^46^.

Self-reported cumulative time spent in organised sports from childhood to adolescence was directly associated with working memory in adolescence among boys but not among girls. This contradicts the results of a previous study that found no sex difference in the associations of self-reported participation in organized sports with cognition^13^. Earlier research has indicated that engaging in sports can improve cognition ^47^. This may be attributed to the greater opportunities for motor learning and cognitively stimulating activities, resulting in beneficial neural adaptations ^48^ and increased possibilities for social interaction and goal-oriented behaviour ^49^. The observed direct association between organized sports and working memory in boys, but not in girls, may be attributed to boys experiencing a more significant acute increase in BDNF following organized sports, including VPA, which is thought to mediate PA-induced cognitive improvements ^24^. The results of our study suggest that participating in organised sports in childhood and adolescence may have beneficial effects on working memory in adolescence among boys. However, the direct association between organised sports and working memory must be interpreted cautiously, as the results did not remain after FDR correction.

Higher levels of device-assessed cumulative LPA from childhood to adolescence were associated with better working memory in adolescence among girls but not among boys. Our findings are consistent with those of Pindus and coworkers ^10^, who observed that higher levels of device-assessed higher-intensity PA were associated with better attentional control in adolescence among girls but not among boys. Our results suggest that even lighter PA could improve cognition in girls. One explanation for our finding might be that device-assessed LPA, particularly in girls, includes talking and socializing with friends, which could enhance cognition ^49^. Activities including socializing may be less important for boys who, in our study, tended to engage more in device-assessed VPA and self-reported organized sports than girls.

The strengths of the present study include a relatively large general population of children followed for eight years until adolescence, valid and reproducible measures of PA, sedentary time, screen time and cognition and the possibility to control for a variety of potential confounders. Moreover, device-assessed and self-reported PA and sedentary time offered us both contextual information on health behaviour and valid information on movement behaviours and gave us a complete picture of PA and sedentary time in childhood and adolescence ^50^. However, our study also includes limitations that need to be considered. The participants’ cognitive performance was only assessed in adolescence, preventing detailed longitudinal analyses. Therefore, it is impossible to determine causality and exclude the possibility of reverse causation, i.e. that individuals with better cognition have less cumulative unsupervised PA and more non-screen-based sedentary time. However, we used the RCPM score at baseline as a covariate in the statistical analyses, which decreases the possibility of reverse causation. In addition, the measures of cumulative PA and sedentary time taken from baseline to the 8-year follow-up may not reflect the possible changes in PA and sedentary time during the period between baseline, 2-year follow-up and 8-year follow-up. Therefore, it is possible that these measures may either overestimate or underestimate the overall exposure to PA and sedentary behaviour in some participants.

## 5. Conclusion

We found that higher levels of self-reported cumulative unsupervised PA and lower levels of self-reported cumulative screen time from childhood to adolescence were associated with poorer cognition in adolescence. We observed no associations of device-assessed cumulative PA or sedentary time from childhood to adolescence with cognition in adolescence. However, higher levels of device-assessed cumulative light PA from childhood to adolescence were associated with better working memory in adolescence among girls but not among boys, suggesting that the associations of PA and sedentary time with cognition partly depend on sex as well as measurement methods and context of PA and sedentary time. Future studies should assess not only PA and sedentary time but also cognition repeatedly to allow complete statistical analyses on longitudinal associations of PA and sedentary behaviours with cognition and provide evidence on these associations from childhood to adolescence and even adulthood.

## Abbreviations

AUC: Area Under the Curve
FDR: False discovery rate
MET: Metabolic equivalent of task
MVPA: Moderate to vigorous physical activity
LPA: Light physical activity
PA: Physical activity
RCPM: Raven Coloured Progressive Matrices
VPA: Vigorous physical activity

## Acknowledgements

The authors would like to thank all adolescents and their parents who participated in the PANIC study and the research group for their skilful contributions in performing these studies.

## Clinical trial registry number

NCT01803776, ClinicalTrials

## Data availability

The datasets generated during and/or analyzed during the current study are not publicly available due to reasons of sensitivity but are available from the corresponding author Jalanko P., via email at.

## Ethics approval statement

The Research Ethics Committee of the Hospital District of Northern Savo approved the study protocol in 2006 (Statement 69/2006) and 2015 (Statement 422/2015).

## Patient consent statement

Written informed consent was obtained from each child’s parent or caregiver, and every child provided assent to participation

