## Supplemental file for "Associations of physical activity and sedentary time from childhood to adolescence with cognition in adolescence: The PANIC study"

**Affiliations**: [a]Faculty of Sport and Health Sciences, University of Jyväskylä, Jyväskylä, Finland; [b] Helsinki Clinic for Sports and Exercise Medicine, Foundation for Sports and Exercise Medicine, Helsinki, Finland; [c] Institute of Biomedicine, School of Medicine, University of Eastern Finland, Kuopio, Finland [d] Faculty of Medicine, University of Helsinki, Helsinki, Finland; [e] Children's Health and Exercise Research Centre, Public Health and Sport Sciences, University of Exeter Medical School, University of Exeter, Exeter, United Kingdom; [f] Institute of Clinical Medicine, Department of Medicine, University of Eastern Finland, Kuopio, Finland; [g] Department of Medicine, Wellbeing Services County of Central Finland, Jyväskylä, Finland; [h] Department of Clinical Physiology and Nuclear Imaging, University of Eastern Finland and Kuopio University Hospital, Kuopio, Finland; [i] Foundation for Research in Health Exercise and Nutrition, Kuopio Research Institute of Exercise Medicine, Kuopio, Finland.

Supplementary Table 1. Associations of self-reported cumulative physical activity, non-screen-based sedentary time and screen time with measures of cognition in 136 boys

| | **Detection Test (reaction time)** | | **Identification Test (reaction time)** | | **One Back Task (reaction time)** | | **Two Back Task (reaction time)** | | **Identification Test (accuracy)** | | **One Back Task (accuracy)** | | **Two Back Task (accuracy)** | | **Continuous Paired Associate Learning Test (errors)** | | **Overall cognition** | |
|---|---|---|---|---|---|---|---|---|---|---|---|---|---|---|---|---|---|---|
| | **β (95% CI)** | **p** | **β (95% CI)** | **p** | **β (95% CI)** | **p** | **β (95% CI)** | **p** | **β (95% CI)** | **p** | **β (95% CI)** | **p** | **β (95% CI)** | **p** | **β (95% CI)** | **p** | **β (95% CI)** | **p** |
| **Total PA** | -0.125 (-0.295, 0.045 | 0.149 | -0.044 (-0.217, 0.129) | 0.617 | -0.068 (-0.239, 0.104) | 0.437 | 0.011 (-0.161, 0.185) | 0.894 | -0.042 (-0.217, 0.133) | 0.635 | 0.149 (-0.018, 0.315) | 0.080 | -0.115 (-0.281, 0.050) | 0.171 | 0.080 (-0.083, 0.244) | 0.333 | -0.026 (-0.189, 0.137) | 0.753 |
| **All organised exercise** | -0.130 (-0.298, 0.038) | 0.127 | -0.111 (-0.281, 0.059) | 0.199 | -0.150 (-0.317, 0.018) | 0.080 | -0.047 (-0.217, 0.124) | 0.589 | -0.057 (-0.229, 0.115) | 0.513 | 0.074 (-0.092, 0.239) | 0.381 | -0.026 (-0.190 0.139) | 0.757 | 0.073 (-0.089, 0.235) | 0.372 | 0.038 (-0.123, 0.199) | 0.642 |
| **Unsupervised PA** | -0.061 (-0.233, 0.111) | 0.484 | 0.035 (-0.140, 0.209) | 0.695 | 0.029 (-0.144, 0.202 | 0.741 | 0.061 (-0.112, 0.235) | 0.486 | -0.012 (-0.188, 0.163) | 0.889 | 0.147 (-0.020, 0.315) | 0.084 | **-0.182 (-0.346, -0.017)** | **0.031*** | 0.067 (-0.098, 0.232) | 0.423 | -0.087 (-0.251, 0.077) | 0.295 |
| **Organised sports** | -0.130 (-0.298, 0.039) | 0.130 | -0.122 (-0.293, 0.048) | 0.159 | **-0.175 (-0.343, -0.008)** | **0.041*** | -0.067 (-0.238, 0.104) | 0.440 | -0.075 (-0.248, 0.098) | 0.391 | 0.049 (-0.118, 0.216) | 0.560 | -0.008 (-0.173, 0.158) | 0.928 | 0.083 (-0.079, 0.245) | 0.315 | 0.040 (-0.122, 0.202) | 0.624 |
| **Non-screen-based ST** | 0.031 (-0.138, 0.200) | 0.717 | -0.085 (-0.255, 0.085) | 0.324 | -0.049 (-0.218, 0.120) | 0.570 | **-0.181 (-0.349, -0.014)** | **0.034*** | 0.016 (-0.156, 0.188) | 0.856 | -0.127 (-0.292, 0.037) | 0.128 | -0.049 (-0.213, 0.115) | 0.554 | 0.027 (-0.135, 0.189) | 0.741 | 0.029 (-0.132, 0.190) | 0.718 |
| **Screen time** | 0.055 (-0.120, 0.230) | 0.538 | -0.089 (-0.266, 0.087) | 0.319 | -0.069 (-0.244, 0.106) | 0.438 | **-0.193 (-0.367, -0.020)** | **0.030*** | 0.073 (-0.105, 0.251) | 0.417 | 0.041 (-0.131, 0.213) | 0.640 | -0.018 (-0.188, 0.153) | 0.836 | -0.056 (-0.223, 0.112) | 0.511 | 0.136 (-0.029, 0.302) | 0.105 |

*Note:* The data are standardised regression coefficients with corresponding 95% confidence intervals (95% CI) and *P*-values for each factor from linear regression analyses adjusted for age, parental education and Raven's Coloured Progressive Matrices baseline score. **P*- value <0.05; ** *P*-value <0.01; *** *P*-value <0.001. Abbreviations: ST, Sedentary time; PA, physical activity

Supplementary Table 2. Associations of device-assessed cumulative physical activity and sedentary time with measures of cognition in 88 boys

| | **Detection Test (reaction time)** | | **Identification Test (reaction time)** | | **One Back Task (reaction time)** | | **Two Back Task (reaction time)** | | **Identification Test (accuracy)** | | **One Back Task (accuracy)** | | **Two Back Task (accuracy)** | | **Continuous Paired Associate Learning Test (errors)** | | **Overall cognition** | |
|---|---|---|---|---|---|---|---|---|---|---|---|---|---|---|---|---|---|---|
| | **β (95% CI)** | **p** | **β (95% CI)** | **p** | **β (95% CI)** | **p** | **β (95% CI)** | **p** | **β (95% CI)** | **p** | **β (95% CI)** | **p** | **β (95% CI)** | **p** | **β (95% CI)** | **p** | **β (95% CI)** | **p** |
| **TPA** | -0.146 (-0.360, 0.069) | 0.182 | -0.025 (-0.245, 0.196) | 0.826 | -0.066 (-0.282, 0.151) | 0.548 | 0.051 (-0.169, 0.272) | 0.644 | 0.056 (-0.276, 0.164) | 0.615 | 0.095 (-0.115, 0.304) | 0.373 | -0.165 (-0.372, 0.041) | 0.114 | 0.006 (-0.203, 0.215) | 0.957 | -0.038 (-0.249, 0.174) | 0.726 |
| **LPA** | 0.001 (-0.219, 0.221) | 0.993 | 0.012 (-0.212, 0.236) | 0.918 | -0.075 (-0.295, 0.144) | 0.497 | -0.141 (-0.363, 0.081) | 0.209 | -0.026 (-0.249, 0.197) | 0.819 | -0.119 (-0.331, 0.093) | 0.268 | 0.061 (-0.151, 0.273) | 0.567 | 0.150 (-0.059, 0.359 | 0.157 | -0.034 (-0.249, 0.181) | 0.752 |
| **MVPA** | -0.073 (-0.305, 0.159) | 0.532 | -0.046 (-0.282, 0.191) | 0.703 | -0.145 (-0.375, 0.086 | 0.215 | -0.015 (-0.251, 0.221) | 0.899 | -0.096 (-0.331, 0.138) | 0.416 | 0.082 (-0.144, 0.307) | 0.473 | 0.177 (-0.044, 0.398) | 0.115 | -0.109 (-0.331, 0.114) | 0.333 | 0.288 (-0.104, 0.347) | 0.288 |
| **VPA** | -0.020 (-0.241, 0.202) | 0.861 | -0.137 (-0.361, 0.087) | 0.227 | -0.066 (-0.287, 0.156) | 0.557 | -0.006 (-0.231, 0.219) | 0.958 | -0.024 (-0.249, 0.200) | 0.830 | 0.048 (-0.167, 0.264) | 0.655 | 0.116 (-0.096, 0.329) | 0.278 | 0.021 (-0.193, 0.234) | 0.846 | 0.052 (-0.164, 0.269) | 0.633 |
| **ST** | 0.078 (-0.157, 0.312) | 0.512 | 0.017 (-0.222, 0.256) | 0.888 | 0.089 (-0.145, 0.323) | 0.450 | 0.039 (-0.200, 0.277) | 0.747 | 0.024 (-0.214, 0.262) | 0.841 | 0.056 (-0.172, 0.283) | 0.627 | -0.169 (-0.392, 0.054) | 0.135 | -0.039 (-0.265, 0.186) | 0.730 | -0.044 (-0.273, 0.185) | 0.701 |

*Note:* The data are standardised regression coefficients with corresponding 95% confidence intervals (95% CI) and *P*-values for each factor from linear regression analyses adjusted for age, parental education and Raven's Coloured Progressive Matrices baseline score. **P*- value <0.05; ** *P*-value <0.01; *** *P*-value <0.001. Abbreviations: TPA, total physical activity; LPA, light physical activity (> 1.5 and ≤ 4.0 standard METs) ; MVPA, moderate-to-vigorous physical activity (> 4.0 standard METs); VPA, vigorous physical activity (> 7.0 standard METs); ST, Sedentary time (≤ 1.5 standard METs, sleep excluded)

Supplementary Table 3. Associations of self-reported cumulative physical activity, non-screen-based sedentary time and screen time with measures of cognition in 124 girls

| | **Detection Test (reaction time)** | | **Identification Test (reaction time)** | | **One Back Task (reaction time)** | | **Two Back Task (reaction time)** | | **Identification Test (accuracy)** | | **One Back Task (accuracy)** | | **Two Back Task (accuracy)** | | **Continuous Paired Associate Learning Test (errors)** | | **Overall cognition** | |
|---|---|---|---|---|---|---|---|---|---|---|---|---|---|---|---|---|---|---|
| | **β (95% CI)** | **p** | **β (95% CI)** | **p** | **β (95% CI)** | **p** | **β (95% CI)** | **p** | **β (95% CI)** | **p** | **β (95% CI)** | **p** | **β (95% CI)** | **p** | **β (95% CI)** | **p** | **β (95% CI)** | **p** |
| **Total PA** | 0.004 (-0.166, 0.173 | 0.967 | -0.118 (-0.296, 0.060) | 0.191 | 0.005 (-0.177, 0.186) | 0.959 | 0.023 (-0.162, 0.209) | 0.803 | 0.105 (-0.076, 0.285) | 0.253 | 0.005 (-0.172, 0.181) | 0.958 | -0.021 (-0.185, 0.142) | 0.797 | -0.059 (-0.239, 0.120) | 0.514 | 0.030 (-0.140, 0.200) | 0.728 |
| **All organised exercise** | -0.004 (-0.172, 0.163) | 0.961 | -0.133 (-0.309, 0.042) | 0.135 | 0.028 (-0.151, 0.207) | 0.755 | -0.006 (-0.190, 0.177) | 0.946 | 0.143 (-0.034, 0.321) | 0.112 | 0.004 (-0.171, 0.178) | 0.968 | 0.039 (-0.123 0.201) | 0.632 | -0.080 (-0.257, 0.098) | 0.375 | 0.064 (-0.103, 0.232) | 0.449 |
| **Unsupervised PA** | 0.001 (-0.168, 0.170) | 0.991 | -0.068 (-0.246, 0.110) | 0.451 | -0.036 (-0.217, 0.145) | 0.692 | 0.001 (-0.184, 0.186) | 0.990 | 0.020 (-0.161, 0.200) | 0.831 | -0.007 (-0.183, 0.169) | 0.936 | -0.049 (-0.212, 0.114) | 0.553 | -0.015 (-0.194, 0.165) | 0.872 | 0.002 (-0.167, 0.172) | 0.979 |
| **Organised sports** | -0.002 (-0.169, 0.165) | 0.981 | -0.120 (-0.296, 0.055) | 0.178 | 0.027 (-0.153, 0.206) | 0.770 | 0.002 (-0.182, 0.185) | 0.987 | 0.099 (-0.079, 0.278) | 0.272 | 0.015 (-0.160, 0.189) | 0.868 | 0.058 (-0.104, 0.219) | 0.480 | -0.082 (-0.260, 0.095) | 0.362 | 0.059 (-0.109, 0.226) | 0.491 |
| **non-screen-based ST** | -0.078 (-0.245, 0.088) | 0.353 | -0.148 (-0.322, 0.026) | 0.096 | -0.148 (-0.324, 0.029) | 0.101 | -0.157 (-0.337, 0.024) | 0.089 | 0.010 (-0.168, 0.189) | 0.910 | -0.149 (-0.321, 0.023) | 0.089 | -0.048 (-0.209, 0.114) | 0.560 | 0.056 (-0.121, 0.233) | 0.534 | 0.066 (-0.101, 0.233) | 0.439 |
| **Screen time** | -0.131 (-0.296, 0.034) | 0.119 | -0.086 (-0.262, 0.089) | 0.331 | **-0.335 (-0.503, -0.167)** | **<0.001***** | **-0.306 (-0.480, -0.132)** | **<0.001***** | -0.023 (-0.202, 0.155) | 0.797 | 0.039 (-0.135, 0.213) | 0.656 | 0.080 (-0.080, 0.241) | 0.325 | 0.013 (-0.164, 0.190) | 0.885 | **0.246 (0.085 0.407)** | **0.003**** |

*Note:* The data are standardised regression coefficients with corresponding 95% confidence intervals (95% CI) and *P*-values for each factor from linear regression analyses adjusted for age, parental education and Raven's Coloured Progressive Matrices baseline score. **P*- value <0.05; ** *P*-value <0.01; *** *P*-value <0.001. Abbreviations: PA, physical activity; ST, Sedentary time

Supplementary Table 4. Associations of device-assessed cumulative physical activity and sedentary time with measures of cognition in 83 girls

| | **Detection Test (reaction time)** | | **Identification Test (reaction time)** | | **One Back Task (reaction time)** | | **Two Back Task (reaction time)** | | **Identification Test (accuracy)** | | **One Back Task (accuracy)** | | **Two Back Task (accuracy)** | | **Continuous Paired Associate Learning Test (errors)** | | **Overall cognition** | |
|---|---|---|---|---|---|---|---|---|---|---|---|---|---|---|---|---|---|---|
| | **β (95% CI)** | **p** | **β (95% CI)** | **p** | **β (95% CI)** | **p** | **β (95% CI)** | **p** | **β (95% CI)** | **p** | **β (95% CI)** | **p** | **β (95% CI)** | **p** | **β (95% CI)** | **p** | **β (95% CI)** | **p** |
| **TPA** | -0.018 (-0.224, 0.187) | 0.860 | -0.181 (-0.397, 0.035) | 0.099 | 0.038 (-0.183, 0.260) | 0.732 | 0.045 (-0.181, 0.272) | 0.691 | 0.177 (-0.046, 0.401) | 0.118 | -0.109 (-0.327, 0.109) | 0.322 | -0.055 (-0.261, 0.150) | 0.595 | -0.149 (-0.370, 0.071) | 0.181 | 0.035 (-0.171, 0.241) | 0.736 |
| **LPA** | 0.008 (-0.199, 0.214) | 0.940 | 0.127 (-0.091, 0.345) | 0.250 | 0.118 (-0.102, 0.339) | 0.289 | 0.182 (-0.042, 0.405) | 0.110 | -0.063 (-0.290, 0.164) | 0.582 | **0.269 (0.057, 0.480)** | **0.014*** | -0.056 (-0.262, 0.150) | 0.591 | 0.076 (-0.147, 0.299 | 0.500 | -0.106 (-0.311, 0.099) | 0.308 |
| **MVPA** | 0.026 (-0.182, 0.234) | 0.804 | -0.015 (-0.237, 0.206) | 0.890 | -0.098 (-0.321, 0.124) | 0.383 | -0.143 (-0.370, 0.083) | 0.211 | 0.215 (-0.009, 0.439) | 0.060 | 0.038 (-0.184, 0.259) | 0.736 | -0.021 (-0.229, 0.187) | 0.844 | -0.008 (-0.233, 0.217) | 0.943 | 0.087 (-0.120, 0.294) | 0.405 |
| **VPA** | -0.002 (-0.207, 0.203) | 0.983 | 0.026 (-0.193, 0.246) | 0.811 | -0.025 (-0.246, 0.196) | 0.824 | -0.046 (-0.272, 0.180) | 0.687 | 0.210 (-0.012, 0.431) | 0.063 | 0.071 (-0.148, 0.289) | 0.522 | -0.022 (-0.227, 0.184) | 0.834 | 0.077 (-0.144, 0.299) | 0.489 | 0.034 (-0.171, 0.239) | 0.741 |
| **ST** | -0.067 (-0.278, 0.144) | 0.526 | -0.135 (-0.359 0.088) | 0.232 | -0.123 (-0.349, 0.103) | 0.282 | -0.141 (-0.372, 0.090) | 0.227 | -0.022 (-0.255, 0.212) | 0.853 | -0.159 (-0.381, 0.064) | 0.161 | 0.045 (-0.167, 0.256) | 0.675 | -0.130 (-0.357, 0.097) | 0.257 | 0.142 (-0.067, 0.351) | 0.180 |

*Note:* The data are standardised regression coefficients with corresponding 95% confidence intervals (95% CI) and *P*-values for each factor from linear regression analyses adjusted for age, parental education and Raven's Coloured Progressive Matrices baseline score. **P*- value <0.05; ** *P*-value <0.01; *** *P*-value <0.001. Abbreviations: TPA; total physical activity; LPA, light physical activity (> 1.5 and ≤ 4.0 standard METs) ; MVPA, moderate-to-vigorous physical activity (> 4.0 standard METs); VPA, vigorous physical activity (> 7.0 standard METs); ST, Sedentary time (≤ 1.5 standard METs, sleep excluded).

Supplementary Table 5. Sex interactions between device-assessed cumulative physical activity and sedentary time and measures of cognition (N=171)

| | **Detection Test (reaction time)** | **Identification Test (reaction time)** | **One Back Task (reaction time)** | **Two Back Task (reaction time)** | **Identification Test (accuracy)** | **One Back Task (accuracy)** | **Two Back Task (accuracy)** | **Continuous Paired Associate Learning Test (errors)** | **Overall cognition** |
|---|---|---|---|---|---|---|---|---|---|
| | **p** | **p** | **p** | **p** | **p** | **p** | **p** | **p** | **p** |
| **TPA** | 0.698 | 0.325 | 0.390 | 0.750 | 0.205 | 0.205 | 0.781 | 0.272 | 0.716 |
| **LPA** | 0.888 | 0.520 | 0.152 | **0.048** | 0.934 | **0.025** | 0.327 | 0.221 | 0.660 |
| **MVPA** | 0.662 | 0.935 | 0.986 | 0.253 | **0.047** | 0.633 | 0.219 | 0.797 | 0.861 |
| **VPA** | 0.951 | 0.478 | 0.923 | 0.638 | 0.117 | 0.909 | 0.434 | 0.869 | 0.863 |
| **ST** | 0.337 | 0.371 | 0.117 | 0.313 | 0.582 | 0.340 | 0.105 | 0.905 | 0.240 |

Adjusted for sex, age, parental education and Raven's Coloured Progressive Matrices baseline score. **P*- value <0.05; ** *P*-value <0.01; *** *P*-value <0.001. Abbreviations: TPA, total physical activity; LPA, light physical activity (> 1.5 and ≤ 4.0 standard METs) ; MVPA, moderate-to-vigorous physical activity (> 4.0 standard METs); VPA, vigorous physical activity (> 7.0 standard METs); ST, Sedentary time (≤ 1.5 standard METs, sleep excluded).

Supplementary Table 6. Sex interactions between self-reported cumulative physical activity, non-screen-based sedentary time and screen time and measures of cognition (N=260)

| | Detection Test (reaction time) | Identification Test (reaction time) | One Back Task (reaction time) | Two Back Task (reaction time) | Identification Test (accuracy) | One Back Task (accuracy) | Two Back Task (accuracy) | Continuous Paired Associate Learning Test (errors) | Overall cognition |
|---|---|---|---|---|---|---|---|---|---|
| | **p** | **p** | **p** | **p** | **p** | **p** | **p** | **p** | **p** |
| **TPA** | 0.560 | 0.578 | 0.540 | 0.924 | 0.323 | 0.378 | 0.810 | 0.200 | 0.742 |
| **All organised exercise** | 0.734 | 0.742 | 0.176 | 0.791 | 0.167 | 0.476 | 0.721 | 0.171 | 0.739 |
| **Unsupervised physical activity** | 0.703 | 0.536 | 0.729 | 0.657 | 0.861 | 0.319 | 0.576 | 0.385 | 0.597 |
| **Organised sports** | 0.687 | 0.962 | 0.118 | 0.628 | 0.240 | 0.603 | 0.706 | 0.134 | 0.822 |
| **non-screen-based ST** | 0.463 | 0.942 | 0.534 | 0.519 | 0.956 | 0.897 | 0.907 | 0.960 | 0.902 |
| **Screen time** | 0.111 | 0.849 | **0.008**** | 0.204 | 0.404 | 0.770 | 0.318 | 0.613 | 0.262 |

Adjusted for sex, age, parental education and Raven's Coloured Progressive Matrices baseline score. **P*- value <0.05; ** *P*-value <0.01; *** *P*-value <0.001. Abbreviations: TPA; total physical activity; ST, Sedentary time (≤ 1.5 standard METs, sleep excluded).
